# Tau-PET and in vivo Braak-staging as a prognostic marker in Alzheimer’s disease

**DOI:** 10.1101/2021.02.04.21250760

**Authors:** Davina Biel, Matthias Brendel, Anna Rubinski, Katharina Buerger, Daniel Janowitz, Martin Dichgans, Nicolai Franzmeier, for the Alzheimer’s Disease Neuroimaging Initiative (ADNI)

**Affiliations:** Institute for Stroke and Dementia Research (ISD), University Hospital, LMU Munich, 81377 Munich,Germany; Department of Nuclear Medicine, University Hospital, LMU Munich, 80336 Munich, Germany; German Center for Neurodegenerative Diseases (DZNE, Munich), Munich, Germany; Munich Cluster for Systems Neurology (SyNergy), Munich, Germany

**Keywords:** Alzheimer’s disease, Braak-staging, tau-PET, amyloid-PET, conversion risk

## Abstract

**INTRODUCTION:** Tau pathology in Alzheimer’s disease tracks clinical status more closely than beta-amyloid. Thus, tau-PET may be a promising prognostic marker for cognitive decline. Here, we systematically compared tau-PET and Braak-staging vs. amyloid-PET as predictors of cognitive decline.

**METHODS:** We included 396 cognitively normal to dementia subjects with ^18^F-Flutemetamol/^18^F-Florbetapir-amyloid-PET, ^18^F-Flortaucipir-tau-PET and ~2-year cognitive assessments. Annual cognitive change rates were calculated via linear-mixed models. We determined global amyloid-PET, global tau-PET, and tau-PET-based Braak-stage (Braak^0^/Braak^I+^/Braak^I-IV+^/Braak^I-VI+^/Braak^atypical+^). In bootstrapped linear regression, we assessed whether tau-PET outperformed amyloid-PET in predicting cognitive decline. Using ANCOVAs, we tested whether later Braak-stage predicted accelerated cognitive decline and determined Braak-stage-specific conversion risk to MCI or dementia.

**RESULTS:** Global tau-PET was a better predictor of cognitive decline than global amyloid-PET (p<0.001). Advanced Braak-stage was associated with faster cognitive decline (p<0.001) and elevated clinical conversion risk.

**DISCUSSION:** Tau-PET and Braak-staging show promise for predicting patient-specific risk of clinical AD progression.

## 1 INTRODUCTION

Beta-amyloid (Aβ) and tau pathology are pathognomonic brain changes of Alzheimer’s disease (AD), ensuing neurodegeneration, cognitive decline and dementia.^1–3^ The development of in vivo Aβ and tau biomarkers has greatly facilitated diagnosing AD,^2,4^ however, reliable prognosis of AD-related cognitive decline remains a critical yet unmet challenge. Tau biomarkers may offer particular potential for individualized risk prediction of AD progression.^5^ While Aβ plaques develop 20-30 years prior to symptom onset,^1,2^ tau pathology emerges much closer to symptom onset as revealed by positron-emission-tomography (PET),^6,7^ biofluid biomarkers,^8^ and post-mortem examinations in AD patients.^9^ Supporting the view that tau is a key determinant of cognitive impairment in AD, PET-based tau assessments show a stronger association with cross-sectional cognition^5^ and retrospective cognitive changes^10^ than amyloid-PET. Similarly, post-mortem studies in AD found that the extent of neurofibrillary tau tangles showed a stronger association with ante-mortem clinical status than Aβ-plaques.^11^ Thus, tau is potentially more informative for forecasting future cognitive decline and clinical progression than Aβ. Also, the spatio-temporal progression of tau pathology follows a relatively stereotypical spreading pattern, which is closely related to disease stage.^12,13^ Specifically, tau emerges first in the entorhinal cortex, before spreading across the temporal lobe, association cortices, and eventually primary sensorimotor and visual cortices, as summarized in the Braak-staging scheme of progressing tau pathology.^12,13^ Thus, tau-PET-based Braak-staging may be clinically useful for patient-specific risk estimation of future cognitive decline and clinical AD progression.

To address this, we systematically tested whether tau-PET and in vivo Braak-staging outperform established amyloid-PET markers as predictors of future cognitive decline and clinical AD progression. To this end, we included 396 subjects ranging from cognitively normal to AD dementia, characterized by baseline ^18^F-Flutemetamol/^18^F-Florbetaben amyloid-PET, ^18^F-Flortaucipir tau-PET, and an average of 2-year follow-up assessments of global cognitive and memory performance. We tested whether i) global measures of tau-PET are a better predictor of future cognitive decline and clinical AD progression than global measures of amyloid-PET and whether ii) advanced tau-PET-assessed Braak-stage predicts faster cognitive decline and risk of clinical AD progression.

## 2 METHODS

### 2.1 Participants

We included 396 participants from the Alzheimer’s Disease Neuroimaging Initiative (ADNI) database. Beyond ADNI inclusion criteria, the current study required availability of amyloid-PET (either 18F-Florbetapir-PET, n=280, or 18F-florbetaben-PET, n=116), together with 18F-Flortaucipir tau-PET and longitudinal cognitive assessments (≥2 examinations), demographics (age, sex, education), and clinical diagnosis. Subjects were categorized by ADNI as cognitively normal (CN, Mini Mental State Examination [MMSE]≥24, Clinical Dementia Rating [CDR]=0, non-depressed), mildly cognitively impaired (MCI, MMSE≥24, CDR=0.5, objective memory-impairment on education adjusted Wechsler Memory Scale II, preserved activities of daily living) or demented (MMSE=20-26, CDR>0.5, NINCDS/ADRDA criteria for probable AD). Note, that ADNI does not include subjects with a baseline MMSE<20, ensuring that despite dementia, cognitive decline can still be assessed in those subjects. For the current study, all baseline PET and cognitive data had to be obtained within a time-window of 6 months. A study flowchart is displayed in Fig. 1A. Ethics approval was obtained by the ADNI investigators from the local ethical committees of all involved sites. Access to all ADNI data was granted to the investigators of the current study after registration to ADNI (https://adni.loni.usc.edu) and compliance with the data usage agreement. The study was conducted in accordance with the Declaration of Helsinki and all study participants provided written informed consent. All work complied with ethical regulations for work with human participants.

**Figure 1:**
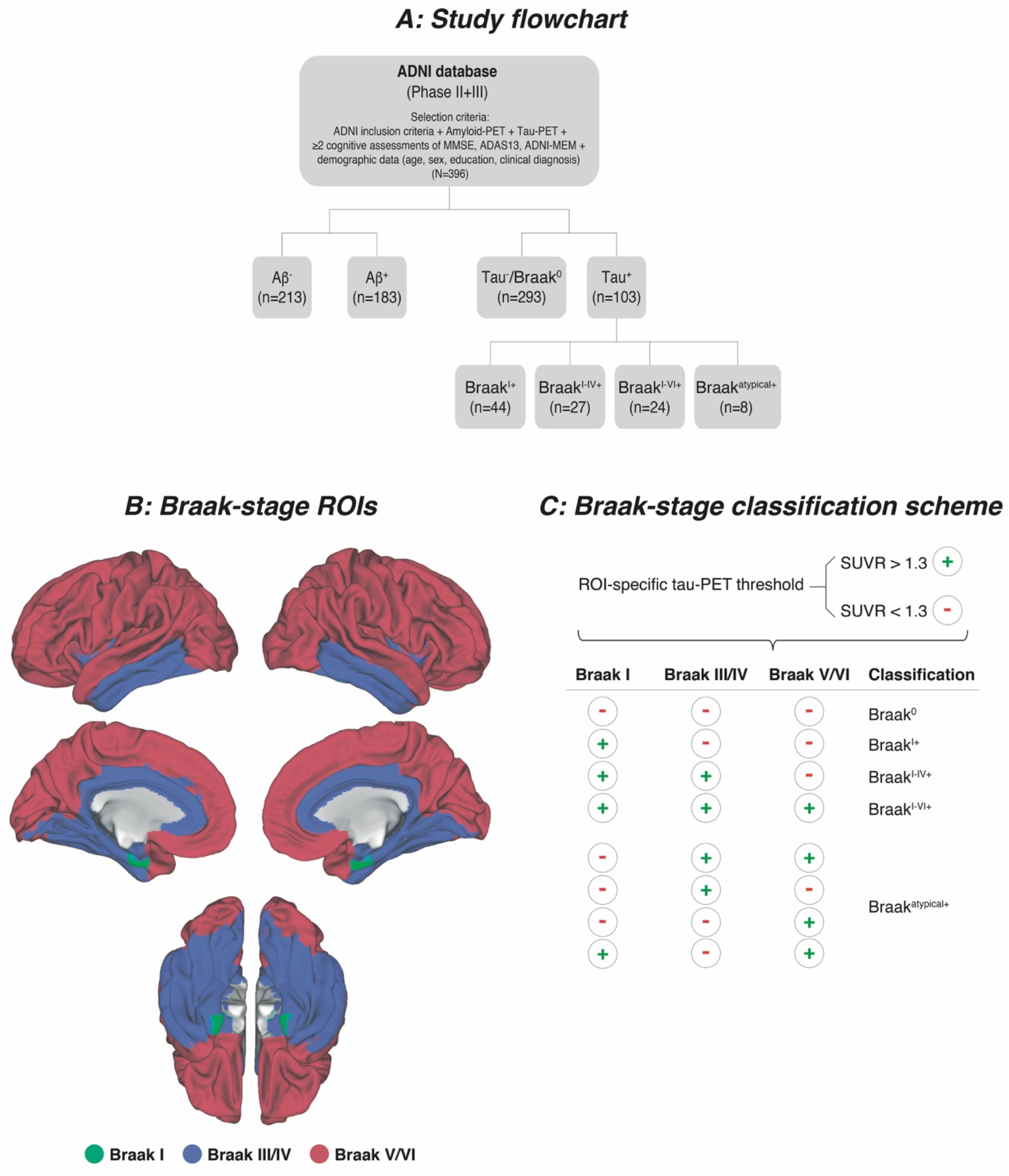
Study flowchart (A), surface rendering of Braak-stage ROIs applied to tau-PET data (B) and tau-PET classification of subjects into Braak-stages (C)

### 2.2 Assessment of cognitive changes

We included longitudinal measures of global cognition and memory. For global cognition, we used the MMSE, i.e. a screening instrument for cognitive deficits that is widely used in clinical routine,^14^ as well as the more extensive Alzheimer’s Disease Assessment Scale Cognition 13-item scale (ADAS13).^15^ Memory performance was assessed using the pre-established ADNI-MEM composite score,^16^ which includes the Rey Auditory Verbal Learning Test, the Alzheimer’s Disease Assessment Scale, the Wechsler Logical Memory Scale I and II, and MMSE word recall. The average cognitive follow-up time was 2.02 years (range=1.00-4.50 years, mean=2.53 cognitive assessments). To determine annual cognitive change rates, we used a pre-established approach,^17,18^ fitting linear mixed models with MMSE, ADAS13, and ADNI-MEM scores as the dependent variable and time (i.e. years from baseline) as the independent variable, controlling for random slope and intercept. From the linear mixed models, we then derived a slope estimate for change in MMSE, ADAS13, ADNI-MEM across time (i.e. change per year) for each subject.

### 2.3 Neuroimaging acquisition and PET preprocessing

3T Structural MRI was obtained using T1-weighted MPRAGE sequences using unified scanning protocols (sequence details can be found on: http://adni.loni.usc.edu/methods/mri-tool/mri-analysis/). Amyloid-PET was recorded within 4×5min blocks 50-70min after ^18^F-Florbetapir injection or 90-110min after ^18^F-florbetaben (FBB) injection. Tau-PET was recorded 75-105min after injection of ^18^F-Flortaucipir within 6⨯5min blocks. For each tracer, recorded timeframes were motion corrected and averaged to obtain a mean image. Structural MRI was preprocessed by ADNI using standard Freesurfer pipelines.

For amyloid-PET, the MRI-derived Freesurfer parcellation^19^ was applied to co-registered PET images to extract global amyloid-PET standardized uptake value ratio (SUVR) values intensity normalized to the whole cerebellum as described previously (see also https://adni.bitbucket.io/reference/docs/UCBERKELEYAV45/ADNI_AV45_Methods_JagustLab_06.25.15.pdf and https://adni.bitbucket.io/reference/docs/UCBERKELEYFBB/UCBerkeley_FBB_Methods_04.11.19.pdf).^20–24^ Subjects were classified as Aβ^+^ when surpassing pre-established SUVR thresholds (^18^F-AV45 SUVR>1.11; ^18^F-florbetaben SUVR>1.2).^23^ To harmonize global amyloid-PET SUVRs across ^18^F-Florbetapir and ^18^F-florbetaben tracers, global SUVR values were transformed to Centiloid (CL) using equations provided by ADNI.^25^

For tau-PET, images were co-registered to structural MRI to extract mean Freesurfer ROI values which were SUVR normalized to the inferior cerebellar grey, following a pre-established approach.^26^ For tau-PET, we obtained global and Braak-stage-ROI-specific (Fig. 1B) tau-PET SUVR scores. For global tau, we averaged SUVRs across cortical Freesurfer ROIs, excluding the cerebellum, hippocampus, thalamus, and basal ganglia (i.e. typical regions of ^18^F-Flortaucipir off-target binding) following a previously described approach.^26^ For Braak-stage specific tau-PET, we applied in vivo Braak-staging that allows application of the post-mortem established Braak tau staging system to tau-PET imaging (see Fig. 1B).^13^ A list of Freesurfer ROIs included within each Braak-stage ROI can be found online (https://adni.bitbucket.io/reference/docs/UCBERKELEYAV1451/UCBERKELEY_AV1451_Methods_Aug2018.pdf). In brief, we obtained tau-PET SUVRs for Braak-stage I, Braak-stage III/IV, and Braak-stage V/VI composite ROIs. Braak-stage II (i.e. hippocampus) was excluded due to ^18^F-Flortaucipir off-target binding in this region.^27^

### 2.4 Braak-staging

Subjects were defined as tau^+^ when at least one Braak-stage ROI surpassed a pre-established cut-off of 1.3 SUVR.^26,28^ For Braak-staging, subjects were classified as Braak I positive (i.e. Braak^I+^, n=44), when only the Braak I ROI (i.e. entorhinal cortex) surpassed a SUVR cut-off of 1.3. Subjects were classified as Braak I-IV positive (i.e. Braak^I-IV+^, n=27) when both Braak I and Braak III/IV ROIs surpassed the SUVR cut-off of 1.3, and as Braak I-VI positive (i.e. Braak^I-VI+^, n=24), when Braak ROIs I, III/IV and V/VI surpassed the 1.3 SUVR threshold. Subjects who deviated from this staging scheme (e.g. for which Braak I was negative, but Braak III/IV was positive) were labeled Braak atypical (i.e. Braak^atypical+^, n=8). Subjects were classified as Braak^0^/tau^-^ (n=293), when all Braak ROIs had an SUVR below 1.3 (Fig. 1A). The Braak-staging scheme is illustrated in Fig. 1C. Note that exploratorily altering the tau-PET threshold between 1.2-1.4 yielded congruent results with those presented in the manuscript.

### 2.5 Statistical analysis

Differences between diagnostic groups were assessed using ANOVAs for continuous and chi-squared (χ^2^) tests for categorial data. To test amyloid-PET and tau-PET as predictors of longitudinal cognitive change rates, we performed linear regression, using annual cognitive change rates (i.e. MMSE, ADAS13, and ADNI-MEM), as dependent variables, and baseline PET (global amyloid-PET SUVR transformed to CL vs. global tau-PET SUVR) as independent variables. Regression models were controlled for age, sex, education, diagnosis (CN, MCI, dementia), and baseline performance on the respective cognitive test (i.e. MMSE, ADAS13, or ADNI-MEM). When assessing global tau-PET SUVR as a predictor of longitudinal cognitive changes, we ran additional models controlling also for global amyloid-PET (i.e. CL), to ensure that tau-PET explained additional variance in cognitive decline when controlling for amyloid-PET. To determine the variance that amyloid-PET (i.e. CL) or tau-PET explained in longitudinal cognitive changes, we calculated partial R^2^ values for either amyloid- or tau-PET as predictors of cognitive changes. In order to assess whether tau-PET was a better predictor of future cognitive changes than amyloid-PET, we performed bootstrapping, repeating the above described regression models on 1000 bootstrapped samples and compared confidence intervals (CI) of amyloid-PET and tau-PET of the resulting partial R^2^ values using paired t-tests. Next, we tested whether more advanced Braak-stage was associated with increased risk of cognitive decline. To this end, we ran ANCOVAs, using annual cognitive change rates as dependent variables (i.e. MMSE, ADAS13, and ADNI-MEM) and Braak-stage group (i.e. Braak^0^, Braak^I+^, Braak^I-IV+^, Braak^I-VI+^, Braak^atypical+^) as independent variable, controlling for age, sex, education, diagnosis, global amyloid-PET, and the baseline score of the respective cognitive test. Standardized differences in cognitive decline between Braak-stage groups were determined using Cohen’s d. Further, we exploratorily assessed whether Braak-stage specific associations with cognitive decline were consistent across diagnostic groups, i.e. we repeated the above described analyses stratified by diagnostic groups (CN, MCI, dementia). Lastly, we determined for each Braak-stage group the risk of clinical conversion, defined as the relative risk of a change in diagnosis from CN to MCI/dementia or from MCI to dementia during follow-up. Note that subjects with a baseline diagnosis of dementia were excluded from this analysis, since no further conversion can be diagnosed in these patients. Conversion rates were compared using χ^2^ tests.

All analyses were computed using R statistical software (r-project.org^29^). For each analysis on cognitive measures (MMSE, ADAS13, ADNI-MEM), Bonferroni correction was applied (adjusted alpha level: 0.05/3=0.017). Post-hoc Tukey tests were applied to ANCOVAs.

### 2.6 Data availability statement

All data used in this manuscript are publicly available from the ADNI database (adni.loni.usc.edu) upon registration and compliance with the data use agreement. The data that support the findings of this study are available on reasonable request from the corresponding author.

## 3 RESULTS

The sample included 239 CN, 122 MCI, and 35 demented individuals. From the CN group, 14 converted to MCI during follow-up (i.e. 5.86% conversion rate), with no conversions to dementia observed. In MCI, 14 converted to dementia during follow-up (i.e. 11.48% conversion rate). Descriptive statistics are displayed in table 1.

**Table 1.** Subjects characteristics.

| **Amyloid<sup>+</sup> (n = 183)** | **CN (n = 93)** | **MCI (n = 60)** | **Dementia (n = 30)** | **p-value** |
| --- | --- | --- | --- | --- |
| Age in years | 75.67 (7.25) | 76.16 (6.82) | 76.95 (7.90) | 0.692 |
| Sex (male/female) | 40/53 | 35/25 | 14/16 | 0.175 |
| Years of education | 16.59 (2.30) | 15.78 (2.73) | 16.1 (2.34) | 0.132 |
| MMSE score | 28.83 (1.48)<sup>b,c</sup> | 27.2 (2.32)<sup>a,c</sup> | 22.27 (3.85)<sup>a,b</sup> | <0.001 |
| ADAS13 score | 12.88 (5.27)<sup>b,c</sup> | 19.44 (6.92)<sup>a,c</sup> | 32.50 (9.32)<sup>a,b</sup> | <0.001 |
| ADNI-MEM score | 0.95 (0.56)<sup>b,c</sup> | 0.13 (0.60)<sup>a,c</sup> | -0.73 (0.73)<sup>a,b</sup> | <0.001 |
| Amyloid-PET CL | 62.10 (32.54)<sup>c</sup> | 75.72 (37.44) | 88.30 (36.61)<sup>a</sup> | <0.001 |
| Global Tau-PET SUVR | 1.13 (0.09)<sup>b,c</sup> | 1.22 (0.18)<sup>a,c</sup> | 1.38 (0.36)<sup>a,b</sup> | <0.001 |
| Braak<sup>0</sup>/Braak<sup>I+</sup>/Braak<sup>I-IV+</sup>/Braak<sup>I-VI+</sup>/Braak<sup>atypical+</sup> | 68/16/6/1/2 | 22/14/13/8/3 | 6/4/6/13/1 | <0.001 |
| Mean cognitive follow-up in years | 2.03 (0.68) | 1.88 (0.87) | 1.67 (0.73) | 0.072 |
| **Amyloid<sup>-</sup> (n = 213)** | **CN (n = 146)** | **MCI (n = 62)** | **Dementia (n = 5)** | **p-value** |
| Age in years | 72.76 (6.87)<sup>b</sup> | 75.47 (8.60)<sup>a</sup> | 71.22 (7.45) | 0.045 |
| Sex (female/male) | 60/86 | 39/23 | 3/2 | 0.014 |
| Years of education | 16.95 (2.42)<sup>c</sup> | 16.18 (3.00) | 14 (2.83)<sup>a</sup> | 0.011 |
| MMSE score | 29.21 (0.98)<sup>b,c</sup> | 28.56 (1.72)<sup>a,c</sup> | 24.4 (1.14)<sup>a,b</sup> | <0.001 |
| ADAS13 score | 11.16 (4.29)<sup>b,c</sup> | 16.19 (5.43)<sup>a,c</sup> | 30.07 (4.39)<sup>a,b</sup> | <0.001 |
| ADNI-MEM score | 1.10 (0.60)<sup>b,c</sup> | 0.60 (0.56)<sup>a,c</sup> | -0.61 (0.59)<sup>a,b</sup> | <0.001 |
| Amyloid-PET CL | 5.60 (8.82)<sup>b</sup> | -0.72 (12.34)<sup>a</sup> | -3.03 (8.87) | <0.001 |
| Global Tau-PET SUVR | 1.09 (0.07) | 1.08 (0.08) | 1.08 (0.07) | 0.811 |
| Braak<sup>0</sup>/Braak<sup>I+</sup>/Braak<sup>I-IV+</sup>/Braak<sup>I-VI+</sup>/Braak<sup>atypical+</sup> | 137/5/1/2/1 | 56/4/1/0/1 | 4/1/0/0/0 | 0.718 |
| Mean cognitive follow-up in years | 2.18 (0.62) | 2.01 (0.92) | 1.53 (0.58) | 0.056 |

Values are presented as mean (SD)
p-values were derived from ANOVAs for continuous measures and from Chi-squared tests for categorical measures
MMSE = Mini-Mental State Examination; ADAS13 = Alzheimer's disease assessment scale, cognitive subscale; ADNI-MEM = episodic memory composite score
Mean values significantly (p < 0.05, Post-hoc tests) different from—
<sup>a</sup> CN
<sup>b</sup> MCI
<sup>c</sup> Dementia

### 3.1 Global tau-PET is a better predictor of future cognitive decline than global amyloid-PET

First, we tested whether global tau-PET at baseline is a better predictor of future cognitive decline than global amyloid-PET (i.e. CL). In linear regression, higher baseline global amyloid-PET was associated with faster subsequent longitudinal cognitive decline for the MMSE (β=−0.074, T=−3.205, p=0.001, partial R^2^=0.026, Fig. 2A), ADAS13 (β=0.138, T=3.830, p<0.001, partial R^2^=0.037, Fig. 2D), and ADNI-MEM (β=−0.055, T=−2.280, p=0.023, partial R^2^=0.013, Fig. 2G). As expected, higher global tau-PET SUVR was also associated with faster cognitive changes (MMSE: β=−0.175, T=−7.528, p<0.001, partial R^2^=0.127, Fig. 2B; ADAS13: β=0.237, T=6.329, p<0.001, partial R^2^=0.093, Fig.2E; ADNI-MEM: β=−0.100, T=−3.927, p<0.001, partial R^2^=0.038, Fig. 2H). The associations between global tau-PET and cognitive decline remained consistent when additionally controlling for global amyloid-PET (MMSE: β=−0.167, T=−6.791, p<0.001, partial R^2^=0.106; ADAS13: β=0.211, T=5.371, p<0.001, partial R^2^=0.069; ADNI-MEM: β=−0.091, T=−3.354, p<0.001, partial R^2^=0.028). Notably, the variance explained (i.e. R^2^) in cognitive decline was higher for global tau-PET compared to global amyloid-PET. To statistically compare the predictive accuracy of amyloid-PET vs. tau-PET, we performed bootstrapping, confirming higher accuracy of tau-PET vs. amyloid-PET in predicting future cognitive decline, i.e. bootstrapped distributions of partial R^2^ values were higher for global tau-PET than for global amyloid-PET (MMSE: T=63.476, p<0.001, Cohen’s d=2.38, Fig. 2C; ADAS13: T=47.645, p<0.001, Cohen’s d=1.76, Fig. 2F; ADNI-MEM: T=50.955, p<0.001, Cohen’s d=1.94, Fig. 2I). In addition, the 95%CIs of the bootstrapped R^2^ distributions did not overlap for MMSE (amyloid-PET: CI=0.026-0.028, mean=0.027; tau-PET: CI=0.126-0.133, mean=0.130), ADAS13 (amyloid-PET: CI=0.041-0.043, mean=0.042; tau-PET: CI=0.100-0.105, mean=0.103), or ADNI-MEM (amyloid-PET: CI=0.029-0.031, mean=0.030; tau-PET: CI=0.075-0.078, mean=0.077), providing non-parametric support of a significant difference between amyloid-PET and tau-PET-derived partial R^2^ distributions.

**Figure 2:**
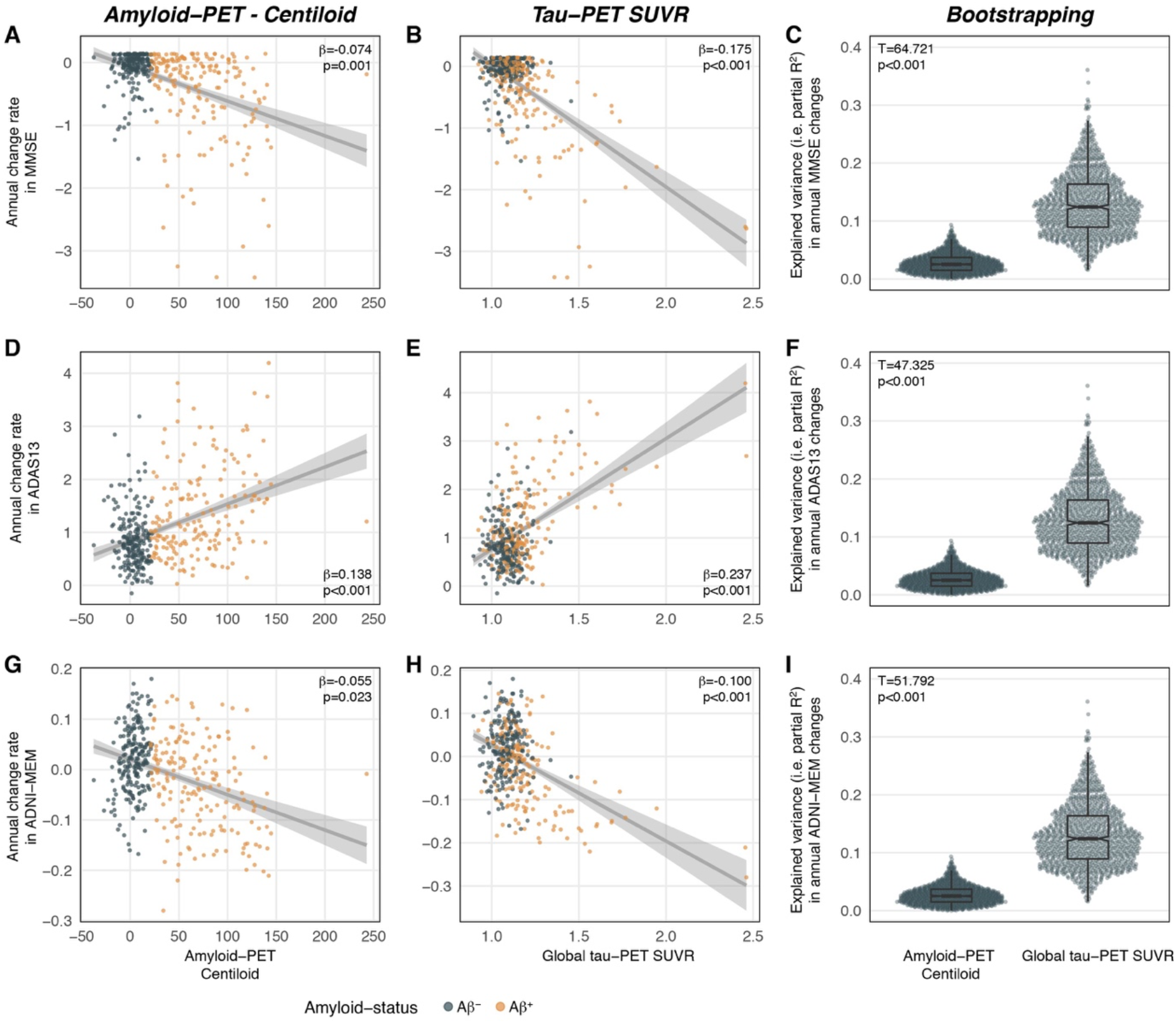
Scatterplot illustrating the association between baseline global amyloid-PET (i.e. Centiloid), global tau-PET SUVRs and annual cognitive changes of the Mini Mental State Examination (MMSE; A+B), Alzheimer’s Disease Assessment Scale Cognition 13-item scale (ADAS13; D+E), and ADNI-MEM (G+H). Standardized beta-values were derived from linear regression controlling for age, sex, education, clinical diagnosis, and the baseline score of the respective cognitive test. Bootstrapping analysis with 1000 iterations (C+F+I) revealed that bootstrapped distributions of partial R^2^ values (i.e. explained variance in cognitive changes) were higher for global tau-PET than for global amyloid-PET.

### 3.2 Advanced Braak-stage is associated with faster cognitive decline

Next, we tested whether more advanced Braak-stage at baseline was associated with faster subsequent cognitive decline using ANCOVAs. We found the expected association between more advanced Braak-stage and faster subsequent cognitive decline consistently for MMSE (F[4,384]=306.099, p<0.001, Fig. 3A), ADAS13 (F[4,380]=107.178, p<0.001, Fig. 3B), and ADNI-MEM (F[4,382]=169.376, p<0.001, Fig. 3C), controlling for age, sex, education, diagnosis, global amyloid-PET, and the baseline score of the respective test. Post-Hoc Tukey tests confirmed significant differences between all sequential Braak-stage groups (all p<0.05). In brief, Braak^0^ subjects showed slowest annual cognitive changes, whereas rates of cognitive decline gradually increased across advancing Braak-stage, with fastest change in Braak^I-VI+^ individuals (Fig. 3). Braak-stage-specific annual cognitive change rates are summarized in table 2. Together, advanced Braak-stage increases the likelihood for future cognitive decline. Standardized differences (Cohen’s d) in cognitive decline between Braak-stage groups are shown in supplementary table 1. Again, this finding supports a close link between advanced tau pathology and the progression of cognitive decline.

**Table 2.**
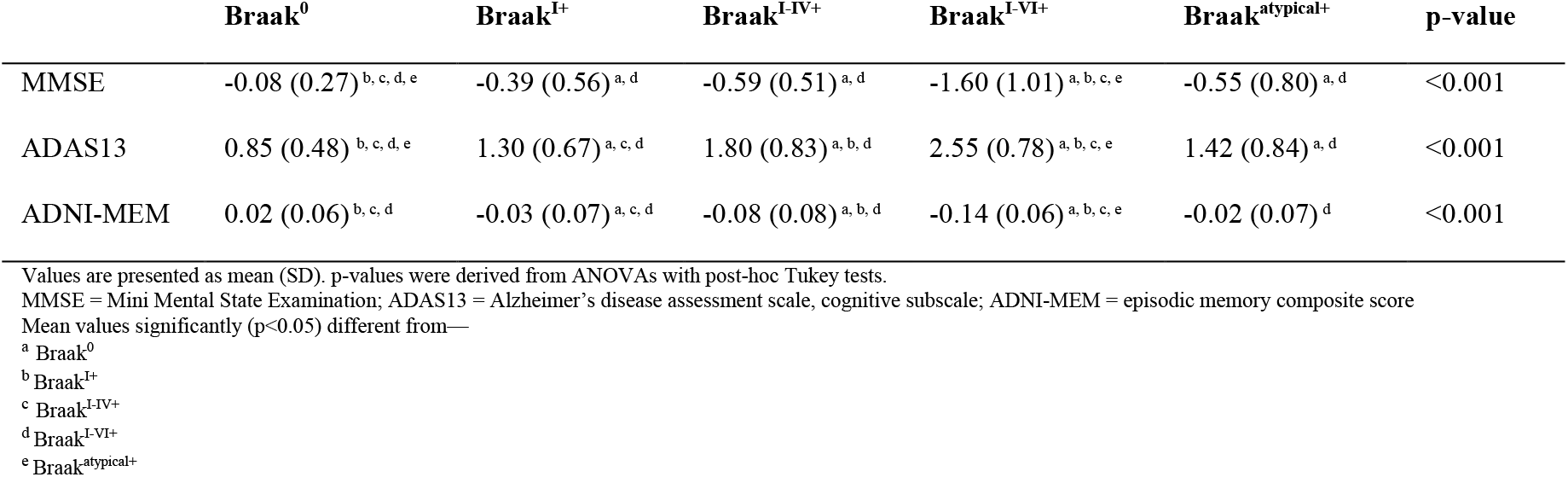
Annual changes in cognition across Braak-stages.

|  | **Braak<sup>0</sup>** | **Braak<sup>I+</sup>** | **Braak<sup>I-IV+</sup>** | **Braak<sup>I-VI+</sup>** | **Braak<sup>atypical+</sup>** | **p-value** |
| --- | --- | --- | --- | --- | --- | --- |
| MMSE | -0.08 (0.27)<sup>b, c, d, e</sup> | -0.39 (0.56)<sup>a, d</sup> | -0.59 (0.51)<sup>a, d</sup> | -1.60 (1.01)<sup>a, b, c, e</sup> | -0.55 (0.80)<sup>a, d</sup> | <0.001 |
| ADAS13 | 0.85 (0.48)<sup>b, c, d, e</sup> | 1.30 (0.67)<sup>a, c, d</sup> | 1.80 (0.83)<sup>a, b, d</sup> | 2.55 (0.78)<sup>a, b, c, e</sup> | 1.42 (0.84)<sup>a, d</sup> | <0.001 |
| ADNI-MEM | 0.02 (0.06)<sup>b, c, d</sup> | -0.03 (0.07)<sup>a, c, d</sup> | -0.08 (0.08)<sup>a, b, d</sup> | -0.14 (0.06)<sup>a, b, c, e</sup> | -0.02 (0.07)<sup>d</sup> | <0.001 |

Values are presented as mean (SD). p-values were derived from ANOVAs with post-hoc Tukey tests.
MMSE = Mini Mental State Examination; ADAS13 = Alzheimer's disease assessment scale, cognitive subscale; ADNI-MEM = episodic memory composite score
Mean values significantly (p<0.05) different from—
<sup>a</sup> Braak<sup>0</sup>
<sup>b</sup> Braak<sup>I+</sup>
<sup>c</sup> Braak<sup>I-IV+</sup>
<sup>d</sup> Braak<sup>I-VI+</sup>
<sup>e</sup> Braak<sup>atypical+</sup>

**Figure 3.**
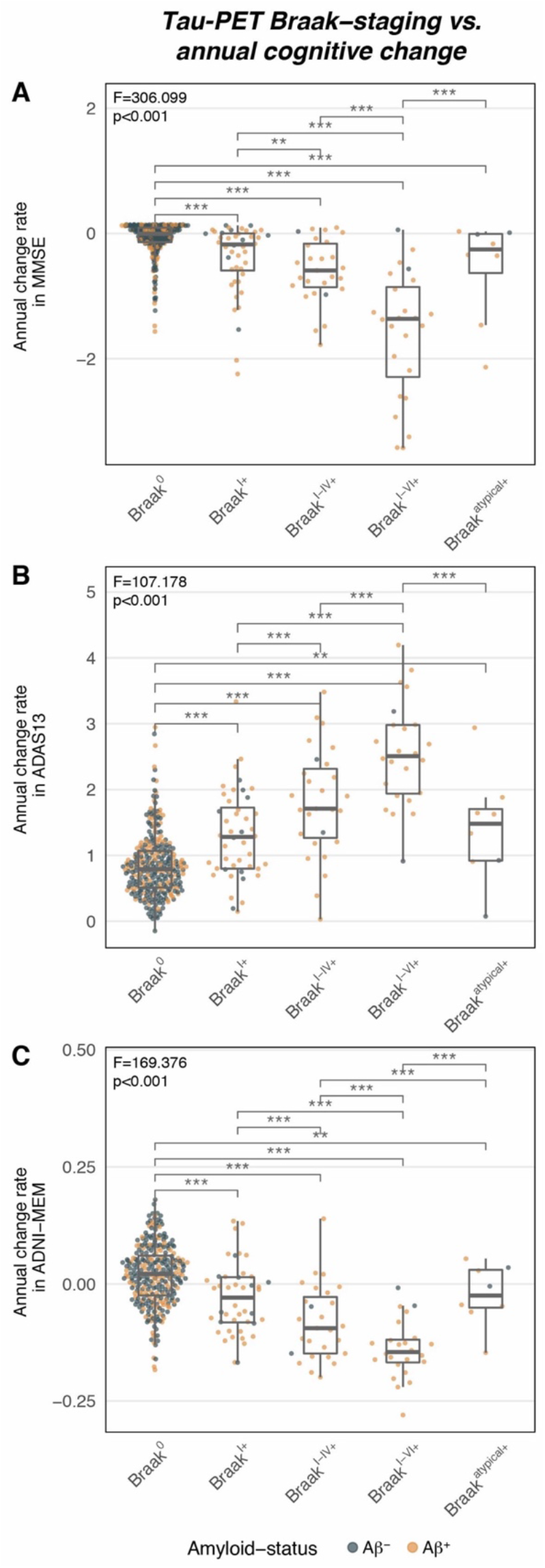
Tau-PET-based Braak-staging versus annual cognitive change rates for the Mini Mental State Examination (MMSE; A), the Alzheimer’s Disease Assessment Scale Cognition 13-item scale (ADAS13; B), and the ADNI-MEM score (C). Statistics were derived from ANCOVA models controlling for age, sex, education, clinical diagnosis, global amyloid-PET (Centiloid), and the baseline score of the respective cognitive test. Post-Hoc Tukey tests were used in order to determine differences in cognitive changes between Braak-stage groups; **= p<0.01, ***=p<0.001.

To test whether the observed associations between Braak-stage and cognitive decline are consistent across diagnostic groups, the above described analyses were repeated stratified by diagnostic group (i.e. CN, MCI, dementia), controlling for age, sex, education, global amyloid-PET, and baseline cognition. Again, associations between advanced Braak-stage and faster subsequent cognitive decline were observed for CN (MMSE: F[4,229]=8.308, p<0.001; ADAS: F[4,227]=11.670, p<0.001; ADNI-MEM: F[4,227]=8.507, p<0.001), MCI (MMSE: F[4,112]=46.253, p<0.001; ADAS: F[4,110]=16.884, p<0.001; ADNI-MEM: F[4,112]=36.690, p<0.001), and dementia (MMSE: F[4,25]=13.993, p<0.001; ADAS: F[4,25]=3.884, p=0.014; ADNI-MEM: F[4,25]=9.158, p<0.001). This supports the view that advancing Braak-stage increases the risk for future cognitive decline across diagnostic groups.

### 3.3 Advanced Braak-stage is associated with higher conversion risk

Lastly, we tested whether Aβ-status or tau-status (i.e. global and Braak-stage specific) were more predictive of future conversion risk from CN to MCI/dementia and from MCI to dementia during follow-up. Note, that 35 participants with baseline dementia diagnosis were excluded from this analysis (see 2.5). Examining the association between baseline Aβ-status or tau-status and clinical conversion during follow-up, Pearson’s χ^2^ test revealed a significant difference between amyloid- and tau-PET positivity and clinical conversion (Aβ-status: χ^2^ [1, N=361]=9.24, p=0.002; tau-status: χ^2^ [1, N=361]=24.962, p<0.001). χ^2^ scores were higher for tau-status compared to Aβ-status, suggesting that tau positivity is more critical for conversion than Aβ positivity. Specifically, Aβ^**+**^ individuals had a conversion risk of 13.07% (n=20/153) vs. 3.85% (n=8/208) in Aβ^**-**^ individuals. In contrast, tau^**+**^ individuals had 21.80% (n=17/78) conversion risk, vs. 3.89% (n=11/283) in tau^**-**^ individuals. Thus, tau positivity is associated with a higher risk for future conversion than Aβ positivity alone. Further supporting this notion, we observed a significant association between Braak-stage (Braak^0^ included) and clinical conversion (X^2^ [4, N=361]=38.925, p<0.001), with a gradual increase in conversion risk across advancing Braak-stage. Specifically, conversion risks were 15.38% (n=6/39) for Braak^I+^, 23.81% (n=5/21) for Braak^I-IV+^, and 45.45% (n=5/11) for Braak^I-VI+^. Individuals classified as Braak^atypical+^ had a conversion risk of 14.29% (n=1/7) (Fig. 4). Again, these results illustrate that spatial expansion of tau pathology is strongly associated with the risk of future cognitive decline, while amyloid-PET is prognostically less conclusive.

**Figure 4.**
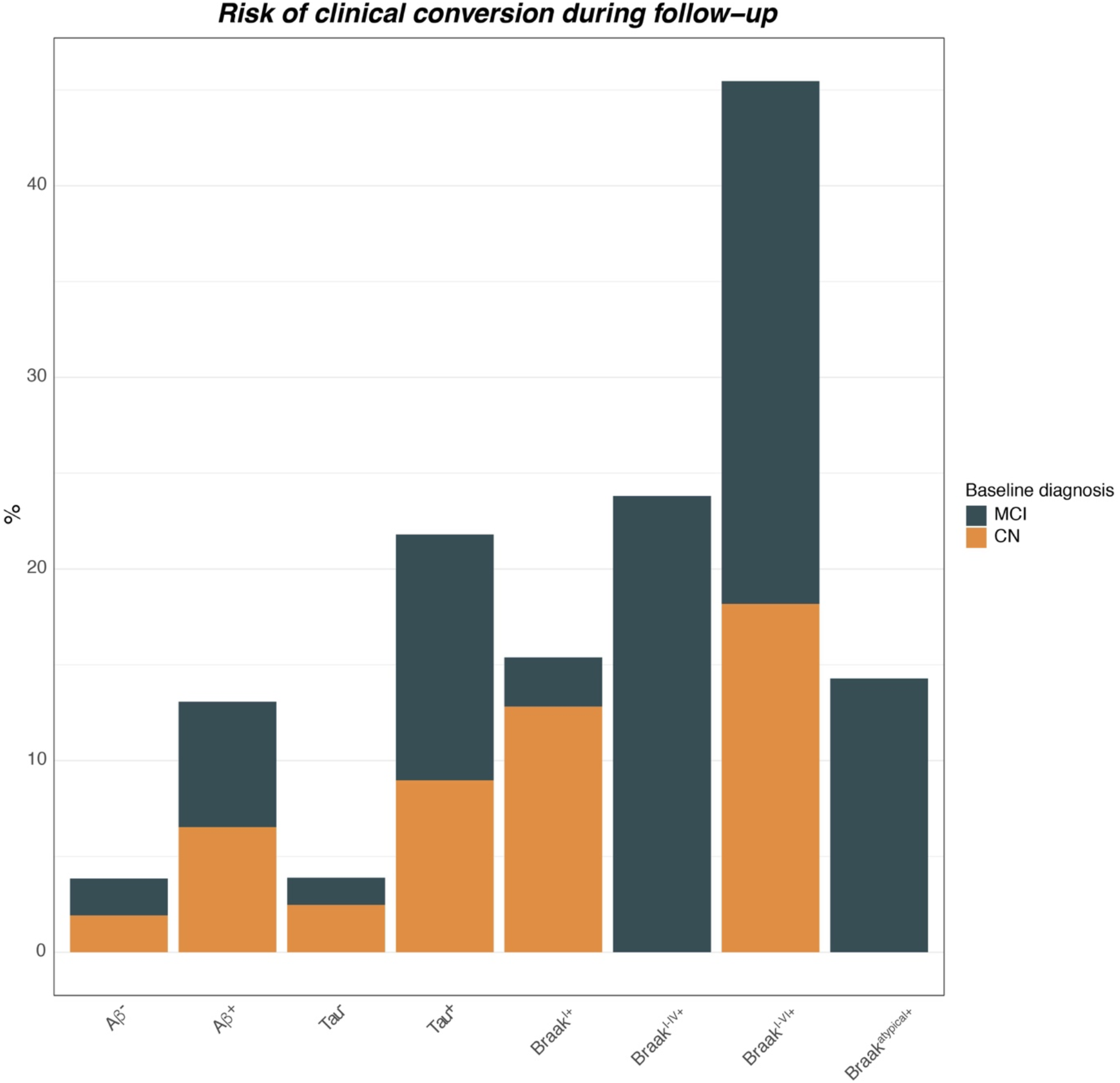
Rates of clinical conversion during follow-up stratified by amyloid-PET positivity, tau-PET positivity and Braak-stage group. Barplots show relative risk of clinical conversion from cognitive normal (CN) to mild cognitive impairment (MCI) or dementia, and from MCI to dementia. Note that subjects with a baseline diagnosis of dementia were excluded from this analysis, since no further diagnostic change can be observed in these participants.

## 4 DISCUSSION

Here, we assessed whether tau-PET outperforms amyloid-PET as a predictor of future cognitive decline and clinical AD progression in older adults with and without cognitive impairment. Supporting this, we found that global tau-PET at baseline explains more variance in subsequent global cognitive and memory decline than amyloid-PET across ~2 years of follow-up. Importantly, the association between global tau-PET and subsequent cognitive changes remained consistent when additionally controlling for baseline amyloid-PET. We further employed PET-based Braak-staging of tau pathology and demonstrate that more advanced Braak-stage is associated with faster subsequent cognitive decline and gradually increasing risk of clinical conversion to MCI or AD dementia, again outperforming the predictive accuracy of amyloid-PET positivity. Together, our results support the view that tau-PET and in vivo Braak-staging may be clinically useful tools for risk prediction of cognitive decline and clinical AD progression.

For our main finding, we show that tau-PET clearly outperforms amyloid-PET in forecasting future cognitive decline. This result pattern is in agreement with several previous studies suggesting a close association between tau accumulation and the development of cognitive deficits in AD.^6,7,9,30^ A recent longitudinal PET-study in preclinical AD showed that increased Aβ-burden mediates tau accumulation which in turn promotes cognitive decline.^30^ This is congruent with the amyloid cascade hypothesis, suggesting that Aβ is the initial trigger of pathological tau in AD^31,32^ preceding symptom onset by decades,^1,2^ while tau is the actual driver of neurodegeneration and cognitive decline.^33^ Therefore, a focus on Aβ biomarkers alone is likely insufficient for reliable prediction of clinical AD trajectories. This has important implications for clinical trial design in AD, since markers of tau pathology could be critical in addition to Aβ-markers for matching progression risk among placebo vs. verum groups. Further, Aβ and tau accumulation show striking differences in their spatial accumulation patterns: While Aβ accumulates rather globally,^34^ tau spreads in a relatively stereotypical spatio-temporal pattern^35^ that is closely associated with clinical status.^13^ By applying Braak-staging to tau-PET, we could confirm that more advanced Braak-stage was associated with gradually accelerated future cognitive decline. Specifically, Braak^0^ individuals (i.e. without evidence of elevated tau pathology) showed slowest annual cognitive changes, whereas Braak^I-VI+^ individuals showed fastest cognitive decline. In line with these findings on continuous rates of cognitive decline, we reported that global tau-PET positivity clearly outperformed global amyloid-PET positivity in predicting future clinical conversion risk (i.e. change in diagnosis) during the ~2-year follow-up period. Braak-stage specific stratification into risk groups further revealed gradually increasing conversion risk at more advanced Braak-stages (e.g. 15.38% for Braak^I+^, 23.81% for Braak^I-IV+^, 45.45% for Braak^I-VI+^). We caution though, that these risk-estimates were based on relatively low clinical conversion rates during the ~2-year follow-up, hence these risk estimates warrant further validation as soon as larger follow-up data become available. Still, our findings support the notion that the spatial expansion of tau pathology holds important information about the risk of future cognitive decline. To our knowledge, this is the first study to systematically compare the accuracy of amyloid-vs. tau-PET and Braak-staging as prognostic markers of cognitive decline, demonstrating that tau-PET is a single promising marker for the prognosis of future cognitive decline and AD progression in clinical settings.

Besides biomarkers of Aβ and tau accumulation, there are numerous other factors that have been associated with cognitive decline and conversion risk, including lifestyle and reserve-related factors (e.g. physical activity, education),^36^ genetic risk (e.g. *APOE4, BIN1, BDNF*, etc.),^37–39^ neuroimmune markers (e.g. TREM2),^40^ and neurodegeneration.^41^ Thus, a combination of lifestyle, genetics as well as clinical and multi-modal biomarkers is likely to yield higher accuracy for predicting cognitive decline and AD progression. Supporting this, we reported previously that a machine-learning model combining structural MRI, FDG-PET, amyloid-PET, and fluid biomarkers for predicting cognitive decline in AD patients outperformed models based on single modalities/biomarkers.^42^ However, multi-modal prediction models including multiple PET scans and fluid biomarkers can be complex and require the assessment of multi-level data which can be challenging to acquire in clinical settings. Thus, our findings on tau-PET as a single and highly predictive biomarker for cognitive decline and clinical progression are potentially of high clinical use.

Several caveats should be considered when interpreting our results. First, ^18^F-Flortaucipir shows considerable off-target binding in the hippocampus and basal ganglia, which may confound the assessment of tau pathology.^43^ Therefore, we excluded regions which are known to be affected by off-target binding. However, influences of unspecific binding remain possible, hence our findings await further replication once sufficient data with second-generation tau-PET tracers (i.e. with a better off-target binding profile) are available. Second, we classified PET using pre-established cut-offs, which is of high clinical use but arbitrarily binarizes a continuous biological process (i.e. Aβ or tau accumulation). Although the currently used tau-PET 1.3 SUVR cut-off was selected based on recommendations for tau-PET,^26,28^ we exploratory repeated our analyses by slightly altering the tau-PET SUVR thresholds (e.g. ranging between 1.2-1.4), revealing a consistent result pattern. Still, PET cut-offs may be replaced in the future by more advanced methods such as gaussian-mixed model-based transformation of tau-PET SUVRs to tau positivity probabilities.^44,45^A third limitation relates to the individual spatial variability in tau deposition patterns. Previous work found that the spreading patterns of tau pathology can be spatially heterogeneous across individual patients.^7,44^ To address this, we applied a relatively simple tau staging scheme (i.e. Braak^I+^, Braak^I-IV+^, Braak^I-VI+^, Braak^atypical^) that does not take into account asymmetry in tau deposition^7^ or fine-grained regional differences in the distribution of tau pathology. Only ~8% of subjects deviated from this staging-scheme, suggesting that the currently employed Braak-staging system is applicable to a majority of typical AD patients that are included in ADNI. Still, we caution that this Braak-staging scheme may not be applicable to patients with atypical AD, characterized by highly heterogeneous tau deposition patterns. Thus, further studies are necessary to determine the predictive accuracy of tau-PET for future cognitive decline in these rare atypical AD cases.^7^ Finally, PET imaging comes with high costs, radioactive burden, and may not be available for each patient or within each country. Therefore, current investigations of plasma tau markers^46^ are of high clinical importance for widespread screening for tau pathology. Plasma screening may be used to select subjects eligible for tau-PET and in vivo Braak-staging, which may allow more accurate risk prediction than single plasma-derived tau measures.

Together, we show that tau-PET outperforms amyloid-PET in predicting cognitive decline and clinical AD progression, supporting the notion that tau pathology is a key driver of cognitive decline in AD.^47^ Importantly, we found that regional tau staging allows accurate risk estimation of future cognitive changes, which can be critical to stratify risk groups for clinical trials. From a clinical perspective, our findings suggest that in vivo tau-PET-based Braak-staging may be a valuable tool to identify subjects at imminent risk of cognitive decline.

## Acknowledgements/Funding

The study was funded by grants from the LMU (FöFoLe, 1032, awarded to NF), the Hertie foundation for clinical neurosciences (awarded to NF), the SyNergy excellence cluster (EXC 2145/ID 390857198) and the German Research Foundation (DFG, INST 409/193-1 FUGG).

## Contributions

D.B.: study concept and design, data processing, statistical analysis, interpretation of the results, and writing the manuscript. M.B.: critical revision of the manuscript. A.R.: critical revision of the manuscript. K.B.: critical revision of the manuscript. D.J.: critical revision of the manuscript. M.D.: critical revision of the manuscript. N.F.: study concept and design, data processing, statistical analysis, interpretation of the results, and writing the manuscript. ADNI provided all data used for this study.

## SUPPLEMENTARY

**Table 1.** Effect sizes between Braak-stage groups and cognitive decline.

|  | MMSE | ADAS13 | ADNI-MEM |
| --- | --- | --- | --- |
| Braak<sup>0</sup> vs. Braak<sup>I+</sup> | -0.95 (large) | 0.88 (large) | -0.76 (medium) |
| Braak<sup>0</sup> vs. Braak<sup>I-IV+</sup> | -1.70 (large) | 1.83 (large) | -1.53 (large) |
| Braak<sup>0</sup> vs. Braak<sup>I-VI+</sup> | -4.00 (large) | 3.31 (large) | -2.45 (large) |
| Braak<sup>0</sup> vs. Braak<sup>atypical+</sup> | -1.58 (large) | 1.14 (large) | -0.65 (medium) |
| Braak<sup>I+</sup> vs. Braak<sup>I-IV+</sup> | 0.37 (small) | -0.69 (medium) | 0.67 (medium) |
| Braak<sup>I+</sup> vs. Braak<sup>I-VI+</sup> | 1.61 (large) | -1.75 (large) | 1.54 (large) |
| Braak<sup>I+</sup> vs. Braak<sup>atypical+</sup> | -0.27 (small) | 0.17 (negligible) | 0.11 (negligible) |
| Braak<sup>I-IV+</sup> vs. Braak<sup>I-VI+</sup> | 1.28 (large) | -0.92 (large) | 0.80 (medium) |
| Braak<sup>I-IV+</sup> vs. Braak<sup>atypical+</sup> | 0.07 (negligible) | -0.46 (small) | 0.77 (medium) |
| Braak<sup>I-VI+</sup> vs. Braak<sup>atypical+</sup> | 1.09 (large) | -1.42 (large) | 1.87 (large) |

Effect sizes were calculated using Cohen's d.
MMSE = Mini Mental State Examination; ADAS13 = Alzheimer's disease assessment scale, cognitive subscale; ADNI-MEM = episodic memory composite score

## REFERENCES

1. Fleisher AS, Chen K, Quiroz YT, et al. Associations between biomarkers and age in the presenilin 1 E280A autosomal dominant Alzheimer disease kindred: a cross-sectional study. JAMA Neurol. 2015;72:316–324.

2. Jack CR, Bennett DA, Blennow K, et al. NIA-AA Research Framework: Toward a biological definition of Alzheimer’s disease. Alzheimers Dement. 2018;14:535–562.

3. Querfurth HW, LaFerla FM. Alzheimer’s disease. N Engl J Med. 2010;362:329–344.

4. Ossenkoppele R, Rabinovici GD, Smith R, et al. Discriminative Accuracy of [18F]flortaucipir Positron Emission Tomography for Alzheimer Disease vs Other Neurodegenerative Disorders. JAMA. 2018;320:1151–1162.

5. Brier MR, Gordon B, Friedrichsen K, et al. Tau and Aβ imaging, CSF measures, and cognition in Alzheimer’s disease. Sci Transl Med. 2016;8:338ra66.

6. Wang L, Benzinger TL, Su Y, et al. Evaluation of Tau Imaging in Staging Alzheimer Disease and Revealing Interactions Between β-Amyloid and Tauopathy. JAMA Neurol. 2016;73:1070–1077.

7. Ossenkoppele R, Schonhaut DR, Schöll M, et al. Tau PET patterns mirror clinical and neuroanatomical variability in Alzheimer’s disease. Brain. 2016;139:1551–1567.

8. Jack CR, Knopman DS, Jagust WJ, et al. Tracking pathophysiological processes in Alzheimer’s disease: an updated hypothetical model of dynamic biomarkers. Lancet Neurol. 2013;12:207– 216.

9. Bennett DA, Schneider JA, Wilson RS, Bienias JL, Arnold SE. Neurofibrillary tangles mediate the association of amyloid load with clinical Alzheimer disease and level of cognitive function. Arch Neurol. 2004;61:378–384.

10. Aschenbrenner AJ, Gordon BA, Benzinger TLS, Morris JC, Hassenstab JJ. Influence of tau PET, amyloid PET, and hippocampal volume on cognition in Alzheimer disease. Neurology. 2018;91:e859–e866.

11. Serrano-Pozo A, Qian J, Muzikansky A, et al. Thal Amyloid Stages Do Not Significantly Impact the Correlation Between Neuropathological Change and Cognition in the Alzheimer Disease Continuum. J Neuropathol Exp Neurol. 2016;75:516–526.

12. Braak H, Braak E. Neuropathological stageing of Alzheimer-related changes. Acta Neuropathol. 1991;82:239–259.

13. Schöll M, Lockhart SN, Schonhaut DR, et al. PET Imaging of Tau Deposition in the Aging Human Brain. Neuron. 2016;89:971–982.

14. Folstein MF, Folstein SE, McHugh PR. “Mini-mental state”. A practical method for grading the cognitive state of patients for the clinician. J Psychiatr Res. 1975;12:189–198.

15. Skinner J, Carvalho JO, Potter GG, et al. The Alzheimer’s Disease Assessment Scale-Cognitive-Plus (ADAS-Cog-Plus): an expansion of the ADAS-Cog to improve responsiveness in MCI. Brain Imaging Behav [online serial]. 2012;6. Accessed at: https://www.ncbi.nlm.nih.gov/pmc/articles/PMC3873823/. Accessed December 7, 2020.

16. Crane PK, Carle A, Gibbons LE, et al. Development and assessment of a composite score for memory in the Alzheimer’s Disease Neuroimaging Initiative (ADNI). Brain Imaging Behav. 2012;6:502–516.

17. Franzmeier N, Suárez-Calvet M, Frontzkowski L, et al. Higher CSF sTREM2 attenuates ApoE4-related risk for cognitive decline and neurodegeneration. Mol Neurodegener. 2020;15:57.

18. Preische O, Schultz SA, Apel A, et al. Serum neurofilament dynamics predicts neurodegeneration and clinical progression in presymptomatic Alzheimer’s disease. Nat Med. 2019;25:277–283.

19. Desikan RS, Ségonne F, Fischl B, et al. An automated labeling system for subdividing the human cerebral cortex on MRI scans into gyral based regions of interest. Neuroimage. 2006;31:968– 980.

20. Franzmeier N, Dewenter A, Frontzkowski L, et al. Patient-centered connectivity-based prediction of tau pathology spread in Alzheimer’s disease. Sci Adv. 2020;6.

21. Franzmeier N, Neitzel J, Rubinski A, et al. Functional brain architecture is associated with the rate of tau accumulation in Alzheimer’s disease. Nature Communications. Nature Publishing Group; 2020;11:347.

22. Franzmeier N, Rubinski A, Neitzel J, et al. Functional connectivity associated with tau levels in ageing, Alzheimer’s, and small vessel disease. Brain. 2019;142:1093–1107.

23. Landau SM, Mintun MA, Joshi AD, et al. Amyloid Deposition, Hypometabolism, and Longitudinal Cognitive Decline. Ann Neurol. 2012;72:578–586.

24. Landau SM, Lu M, Joshi AD, et al. Comparing PET imaging and CSF measurements of Aβ. Ann Neurol. 2013;74:826–836.

25. Klunk WE, Koeppe RA, Price JC, et al. The Centiloid Project: Standardizing quantitative amyloid plaque estimation by PET. Alzheimer’s & Dementia. 2015;11:1-15.e4.

26. Maass A, Landau S, Baker SL, et al. Comparison of multiple tau-PET measures as biomarkers in aging and Alzheimer’s disease. Neuroimage. 2017;157:448–463.

27. Lemoine L, Leuzy A, Chiotis K, Rodriguez-Vieitez E, Nordberg A. Tau positron emission tomography imaging in tauopathies: The added hurdle of off-target binding. Alzheimers Dement (Amst). 2018;10:232–236.

28. Jack CR, Wiste HJ, Weigand SD, et al. Defining imaging biomarker cut-points for brain aging and Alzheimer’s disease. Alzheimers Dement. 2017;13:205–216.

29. R Core Team. R: A language and environment for statistical computing. R Foundation for Statistical Computing, Vienna, Austria. URL https://www.R-project.org/. Vienna, Austria; 2020.

30. Hanseeuw BJ, Betensky RA, Jacobs HIL, et al. Association of Amyloid and Tau With Cognition in Preclinical Alzheimer Disease: A Longitudinal Study. JAMA Neurol. Epub 2019 Jun 3.

31. Hardy J, Selkoe DJ. The amyloid hypothesis of Alzheimer’s disease: progress and problems on the road to therapeutics. Science. 2002;297:353–356.

32. Glenner GG, Wong CW. Alzheimer’s disease and Down’s syndrome: sharing of a unique cerebrovascular amyloid fibril protein. Biochem Biophys Res Commun. 1984;122:1131–1135.

33. La Joie R, Visani AV, Baker SL, et al. Prospective longitudinal atrophy in Alzheimer’s disease correlates with the intensity and topography of baseline tau-PET. Sci Transl Med. 2020;12.

34. Selkoe DJ, Hardy J. The amyloid hypothesis of Alzheimer’s disease at 25 years. EMBO Mol Med. 2016;8:595–608.

35. Braak H, Thal DR, Ghebremedhin E, Del Tredici K. Stages of the pathologic process in Alzheimer disease: age categories from 1 to 100 years. J Neuropathol Exp Neurol. 2011;70:960– 969.

36. Stern Y, Arenaza-Urquijo EM, Bartrés-Faz D, et al. Whitepaper: Defining and investigating cognitive reserve, brain reserve, and brain maintenance. Alzheimers Dement. 2020;16:1305– 1311.

37. Corder EH, Saunders AM, Strittmatter WJ, et al. Gene dose of apolipoprotein E type 4 allele and the risk of Alzheimer’s disease in late onset families. Science. 1993;261:921–923.

38. Franzmeier N, Rubinski A, Neitzel J, Ewers M, Alzheimer’s Disease Neuroimaging Initiative (ADNI). The BIN1 rs744373 SNP is associated with increased tau-PET levels and impaired memory. Nat Commun. 2019;10:1766.

39. Franzmeier N, Ren J, Damm A, et al. The BDNFVal66Met SNP modulates the association between beta-amyloid and hippocampal disconnection in Alzheimer’s disease. Mol Psychiatry. Epub 2019 Mar 21.

40. Ewers M, Franzmeier N, Suárez-Calvet M, et al. Increased soluble TREM2 in cerebrospinal fluid is associated with reduced cognitive and clinical decline in Alzheimer’s disease. Sci Transl Med. 2019;11.

41. Jack CR, Petersen RC, Xu YC, et al. Prediction of AD with MRI-based hippocampal volume in mild cognitive impairment. Neurology. 1999;52:1397–1403.

42. Franzmeier N, Koutsouleris N, Benzinger T, et al. Predicting sporadic Alzheimer’s disease progression via inherited Alzheimer’s disease-informed machine-learning. Alzheimers Dement. 2020;16:501–511.

43. Leuzy A, Chiotis K, Lemoine L, et al. Tau PET imaging in neurodegenerative tauopathies-still a challenge. Mol Psychiatry. 2019;24:1112–1134.

44. Franzmeier N, Dewenter A, Frontzkowski L, et al. Patient-centered connectivity-based prediction of tau pathology spread in Alzheimer’s disease. Sci Adv. 2020;6.

45. Vogel JW, Iturria-Medina Y, Strandberg OT, et al. Spread of pathological tau proteins through communicating neurons in human Alzheimer’s disease. Nat Commun. 2020;11:2612.

46. Palmqvist S, Janelidze S, Quiroz YT, et al. Discriminative Accuracy of Plasma Phospho-tau217 for Alzheimer Disease vs Other Neurodegenerative Disorders. JAMA. 2020;324:772–781.

47. Bejanin A, Schonhaut DR, La Joie R, et al. Tau pathology and neurodegeneration contribute to cognitive impairment in Alzheimer’s disease. Brain. 2017;140:3286–3300.

